# Profiling small RNAs in CRC screening samples such as the widely used fecal immunochemical test, is it possible?

**DOI:** 10.1101/2023.05.03.23289251

**Authors:** Einar Birkeland, Giulio Ferrero, Barbara Pardini, Sinan U. Umu, Sonia Tarallo, Sara Bulfamante, Geir Hoff, Carlo Senore, Trine B Rounge, Alessio Naccarati

## Abstract

Faecal microRNAs represent promising molecules with potential clinical interest as non-invasive diagnostic and prognostic biomarkers for their stability and detectability. Colorectal cancer (CRC) screening based on the fecal immunochemical test (FIT) is an effective tool for prevention of cancer development. However, due to the poor sensitivity of FIT for premalignant lesions, there is a need for implementation of complementary tests. Improving the identification of individuals who would benefit from further investigation with colonoscopy using molecular analysis, such as miRNA profiling of the FIT leftover buffer, would be ideal due to its widespread use.

In the present study, we applied small RNA sequencing to FIT leftover samples collected from two European screening populations. We showed robust detection of miRNA and microbial profiles, which were similar to those obtained from specimens sampled using RNA stabilising buffers and archived fecal samples. Detected miRNAs exhibited differential abundance between CRC and control samples that was consistent between sampling methods, suggesting a promising potential to identify small RNA CRC biomarkers using FIT leftovers. We demonstrated that it is possible to analyse gut miRNAs in FIT leftover samples and envision that these potential biomarkers can complement the FIT in large scale screening settings.

## Introduction

Colorectal cancer (CRC) is the third most common malignancy and the second leading cause of cancer-related deaths(1). The promotion of healthy lifestyles and dietary choices, the development of new strategies for disease management, and the implementation of global screening programs are some of the strategies to reduce CRC morbidity and mortality.

Screening selected age groups at risk is considered the most effective tool to prevent CRC development by detecting early tumor forms and precancerous lesions(2). In many European countries, the first step of CRC screening relies on non-invasive stool-based tests such as fecal immunochemical test (FIT). If the test is positive, patients are invited to visual examinations based on invasive endoscopic methods, such as colonoscopy. The advantage of first-line FIT is the relatively low-cost and ease of execution compared to colonoscopy. The FIT does not require specific preparation or dietary restriction and consequently has high acceptance rates. However, due to its poor sensitivity for premalignant lesions and the burden associated with an excessive number of colonoscopy procedures, different countries adopt thresholds for FIT positivity that are suited to their colonoscopy capacity, in a balancing act between sensitivity and specificity.

This highlights the need for alternative biomarkers to improve CRC screening accuracy. The implementation of complementary tests based on the analysis of the leftover of FIT stool samples could help improve in the identification of those individuals that would benefit from further investigation by colonoscopy. Both observational and experimental evidence point to a role for the gut microbiome in development and progression of CRC (3). We have shown that it is possible to profile the microbiome in FIT leftover samples and archived stool samples (4). However, larger discovery studies are needed to identify clinically valuable biomarkers(5). Small noncoding RNAs (sRNAs), particularly microRNAs (miRNAs), are detectable and stable in stool samples and are emerging as a candidate source of biomarkers for the non-invasive diagnosis of gastrointestinal diseases, including CRC(6). Using small RNA sequencing, we demonstrated the possibility to quantify the levels of both human and microbial sRNAs in human stool samples. Interestingly, the combined use of human and microbial sRNA levels was more efficient than using the two biomarkers alone in classifying CRC patients from colonoscopy-negative control subjects(7).

While we have shown that gut derived miRNAs are potential biomarkers for CRC, little is known about the possibility of measuring them in a screening population by using the FIT buffer leftovers. In this study, carried out in two independent European laboratories, we showed not only the feasibility of small RNA sequencing in FIT samples but we also tested the profiling robustness by comparing sequencing data in FIT samples with feces collected in stabilising buffers and long term archived fecal samples. In addition, we showed that some gut miRNAs differed in abundance between CRC and controls, suggesting a potential for discovering CRC biomarkers.

## Material and Methods

### Cohorts and samples

#### BCSN – FIT

Bowel Cancer Screening in Norway (BCSN) is an ongoing (2012-) randomized trial comparing once-only sigmoidoscopy with repeated FIT tests every second year for up to four rounds. The study is a pilot for the national screening program(8). Stool samples are collected on plastic sticks designed to catch about 10μg stool and then stored in a 2ml buffer (Eiken Chemicals Ltd., Tokyo, Japan). Thirteen FIT samples from anonymized subjects were randomly selected for the purpose of this feasibility study.

#### NORCCAP – stool

The Norwegian Colorectal Cancer Prevention (NORCCAP) screening trial was carried out from 1999 to 2001(9). Participants were asked to bring a fresh frozen stool specimen collected at home less than one week before sigmoidoscopy and to keep in a 20ml vial in their home deep freezer (−20°C) until attendance for flexible sigmoidoscopy. Eleven anonymized stool samples were randomly selected.

#### MITOS - FIT and stool

The Italian biological samples have been collected in the frame of the regular Piedmont Region CRC screening in the Microbiome and MiRNA in Torino Screening project (MITOS). The Piedmont Region screening program invites all residents, aged 59-69 to undergo a single sample biennial FIT (Eiken Chemicals Ltd., Tokyo, Japan). A total of 185 subjects (based on colonoscopy results classified in 22 CRC, 80 advanced adenoma (AA), 30 non-advanced adenoma (nAA), and 53 controls) were included in the present study. Among them, 57 subjects also provided stool samples collected in nucleic acid collection and transport tubes with RNA stabilising solution (Norgen Biotek Corp.) before undergoing colonoscopy.

### small RNA extraction and library preparation

FIT stool samples were obtained from buffer leftovers contained in the original collection device (approx. 1ml). NORCCAP feces was thawed and homogenized in a buffer (Omnigene-GUT, DNAgenotek). For BCSN FIT and NORCCAP stool samples, RNA was extracted from 200µl buffer leftovers and buffer mix, respectively. RNA was purified using phenol-chloroform phase separation and miRNeasy Mini Kit (cat. no. 217004, QIAgen).

For the MITOS cohort, total RNA from stool and FIT leftover samples was extracted using the Stool Total RNA Purification Kit (Norgen Biotek Corp.) as previously described(7).

sRNA transcripts were converted into barcoded cDNA libraries with the NEBNext Multiplex Small RNA Library Prep Set for Illumina following the NEBNext Multiplex Small RNA Library Prep Protocol E7330 (Protocol E7330, New England BioLabs Inc., USA)(7, 10).

The size selection of purified RNA fragments for the MITOS cohort was performed as described in (7). For BCSN and NORCCAP samples the size selection was performed with a cut size optimized to cover RNA molecules from 17 to 47 nt in length. Small RNA libraries were indexed and sequenced on Illumina platforms.

### Bioinformatics and statistical analysis

Quantification of reads mapping to miRBase v22.1 miRNA sequences was performed using the smrnaseq pipeline (https://nf-co.re/smrnaseq) with reads prefiltered using fastp and the *skip_mirdeep* option. Reads unmapped on these annotations were aligned against the human hg38 genome using Bowtie2 with *–very-sensitive-local* option. The human-unmapped reads were mapped against microbial genomes using Kraken 2 (v2.1.2) as described in(7).

Reads mapping to miRNAs were normalized and Differential Expression (DE) analyses were performed using the DESeq2. Differential abundance analysis of reads mapped to microbes was performed with SIAMCAT with default settings. For more details, see Supplementary Methods.

## Results and discussion

### Cohort and alignment statistics

Fecal samples collected with different sampling, storage, and processing procedures were analysed to identify stably expressed miRNAs with a potential to be used as CRC biomarkers (**Figure 1A**). From the MITOS cohort, two sample types were collected for each individual: stool samples collected in RNA-stabilising buffer, and leftover buffer derived from CRC-screening FIT samples. Two sets of anonymized Norwegian samples were also assessed: archived stool samples stored without stabilising buffer from the NORCCAP study, and leftover buffer from Norwegian CRC-screening FIT samples from the BCSN trial.

**Figure 1.**
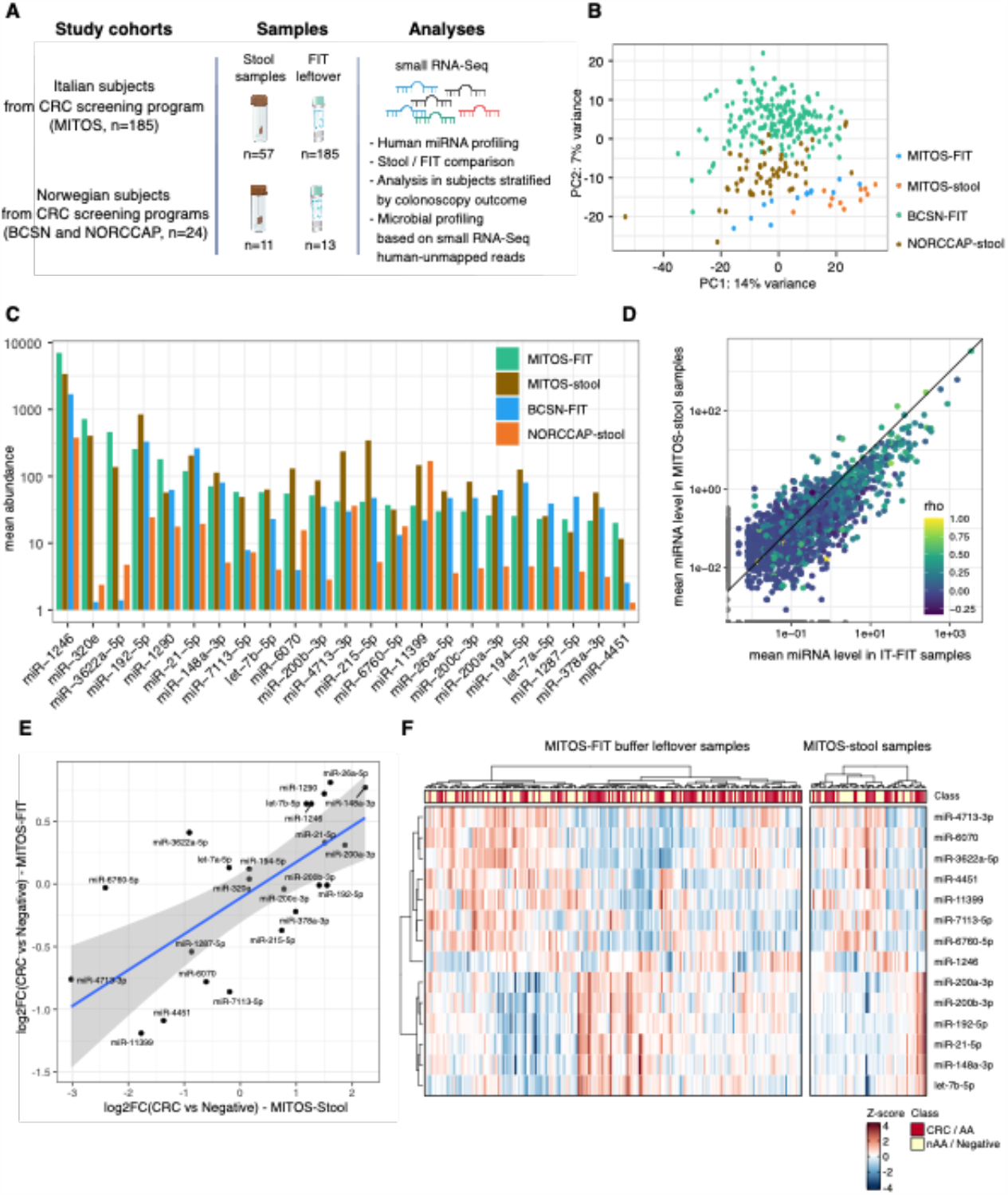
**A**. Graphical representation of the study. **B**. PCA of samples based on mature miRNA read counts. **C**. Bar plot reporting, for each cohort and sample type, the average levels of the miRNAs associated with the highest levels in MITOS FIT-leftover samples. **D**. Mean normalised abundance of miRNAs in paired MITOS-FIT (x-axis) and MITOS-Stool (y-axis) samples. Each point represents a miRNA coloured based on the Spearman correlation coefficient for comparison of paired samples. Only miRNAs detected in at least 15% of each sample type were included in this analysis. **E**. Scatterplot showing the log2FC of expression computed considering MITOS-stool (x-axis) and MITOS-FIT-leftover (y-axis) miRNA levels in samples from CRC patients with respect to those from colonoscopy-negative subjects. **F**. Heatmap of the Z-score-normalised DE miRNA levels in MITOS-stool and MITOS-FIT-leftover samples. Neg, colonoscopy negative subjects; AA, advanced adenoma; nAA, non-advanced adenoma; CRC, colorectal cancer.

As reported in **Supplementary Table 1A** and **B**, a mean of 0.12% of small RNA sequencing reads (range: 0.003-0.062%) were assigned to miRNAs in stool samples, while a mean of 0.15% reads from FIT leftovers were assigned to miRNAs (range:0.03-0.61%). Considering 10 as the minimum number of normalised reads to define a miRNA as detected, on average 63 (range: 32-235) and 41 (range: 16-191) miRNAs were detected in stool and FIT leftover samples, respectively (**Supplementary Table 1A-B**). Still, when accounting for differences in sequencing depth by rarefaction, no differences in the number of miRNAs detected were found between sampling groups (p>0.1). The stool miRNAs detected in the MITOS cohort samples included most of the annotations observed in previous analyses performed on the same samples analysed using a different pipeline(7).

For two FIT leftover samples, the small RNA sequencing was performed on libraries generated from two different amounts of starting material (250 and 400 µl). Comparing the rate of miRNA-mapped reads, similar rates were observed (0.4-0.9% of mapped reads) with an average of 33 miRNAs (range: 31-34) detected in each experiment. No significant differences were observed among the miRNA levels measured in such experiments and, as expected, the levels of detected molecules were significantly correlated (rho=0.43-0.60, p<0.001) (**Supplementary Figure 1A** and **Supplementary Table 1C**). The intra-individual correlation between miRNA levels was higher than among different individuals (**Supplementary Figure 1B**). Taken together all fecal/FIT samples collected in all cohorts, t FIT leftover samples differed systematically from stool samples (see PCA analysis in **Figure 1B)**.

Since the 57 stool samples from the MITOS cohort were collected from the same subjects donating FIT leftover samples, a paired comparative analysis was performed between the miRNA levels measured in the two biospecimens. The analysis was focused on the levels of 23 miRNAs that were consistently identified across both stool and FIT samples (exceeding a median of 10 normalised reads; **Figure 1C** and **Supplementary Table 1D**). These miRNAs correspond well with the most abundant miRNAs in circulation, including miR-1246, -320, -21-5p,-1290,-148a-3p being among the top 10 abundant miRNAs in serum samples(11). As reported in **Supplementary Table 1D**, 57% of these miRNAs were characterised by levels positively correlated (average rho=0.36, p<0.05) between the two biospecimens. Among them, miR-4713-3p, miR-1246, and miR-192-5p were characterised by the highest correlation. All the 23 miRNAs were also detected in the Norwegian cohorts (**Figure 1C)**, despite the different sampling, preservation, and RNA extraction procedures. Moreover, there was a positive correlation between FIT and stool samples for the mean normalised abundance of miRNAs (rho=0.78; **Figure 1D**), which was also found in the Norwegian samples (rho=0.52) (**Figure 1D** and **Supplementary Figure 1C**).

### miRNA differential expression in MITOS stool and FIT samples

DE analysis was performed between MITOS-stool and MITOS-FIT miRNA levels detected in subjects with CRC or AA with respect to colonoscopy-negative subjects. Considering the 23 miRNAs detected in both biofluids, 12 were associated with significantly different levels in stool samples from CRC/AA patients (adj. p<0.05; **Supplementary Table 1E**). Comparing separately AA and CRC patients with colonoscopy-negative subjects, four and eight DE miRNAs were observed, respectively. Two miRNAs (miR-192-5p, and let-7b-5p) were DE in both comparisons. Conversely, four miRNAs were significantly more (let-7b-5p and miR-148a-3p) and less (miR-4451 and miR-11399) abundant in FIT leftover samples of CRC patients with respect to colonoscopy-negative subjects (**Supplementary Table 1F)**. The levels of the 23 miRNAs were characterised by a coherent difference in both MITOS-stool and MITOS-FIT samples (rho=0.69, p<0.001) (**Figure 1E-F**).

Functional analysis of DE miRNA target genes showed the prevalence of terms related to cell cycle regulation and DNA-damage response for the targets of miRNAs with increased levels in CRC patients (**Supplementary Table 1G-H**). Conversely, targets of miRNAs decreasing in patient samples were enriched in processes related to apoptosis, unfolded protein stress response, and immune response (**Supplementary Table 1G-H**).

### Microbial classification/diversity

After the identification of human miRNAs, the remaining reads were aligned against the human genome and the subsequent unmapped reads were investigated for their microbial sRNA content. This approach classified reads in the range of 36-40 nt, of which 38.5% and 37.4% were classified in the MITOS stool and FIT samples, respectively. Given the previous evidence on the concordance between microbial abundances estimated by small RNA sequencing and metagenomic data(7), the human-unmapped sRNA-Seq reads were used to infer the microbial abundance in our data. Overall, FIT samples displayed a higher abundance of taxa belonging to the *Bacteroidetes* phylum, whereas stool samples were dominated by *Firmicutes* (**Figure 2A-B**), with the composition of stool and FIT samples differing significantly (PERMANOVA p < 0.05; **Figure 2C**). This could indicate a differential sensitivity of bacteria to the buffer components in the FIT and NORGEN buffers, where the former has a relatively high concentration of the potent antimicrobial compound sodium azide (12). Still, at the species level, there was concordance between FIT and stool samples (**Figure 2D**). Within the MITOS study subjects, we assessed differences in microbial taxa between colonoscopy-negative and either CRC, AA, or CRC/AA. Only one species - *Filimonas lacunae* - was found to be significantly more abundant in stool samples collected from CRC cases than in controls, with nominal significance also observed in FIT samples. Like for human miRNAs, the direction and magnitude of differences in taxa abundance between CRC cases and colonoscopy-negative subjects were consistent in stool and FIT samples (rho = 0.48, p <0.001).

**Figure 2.**
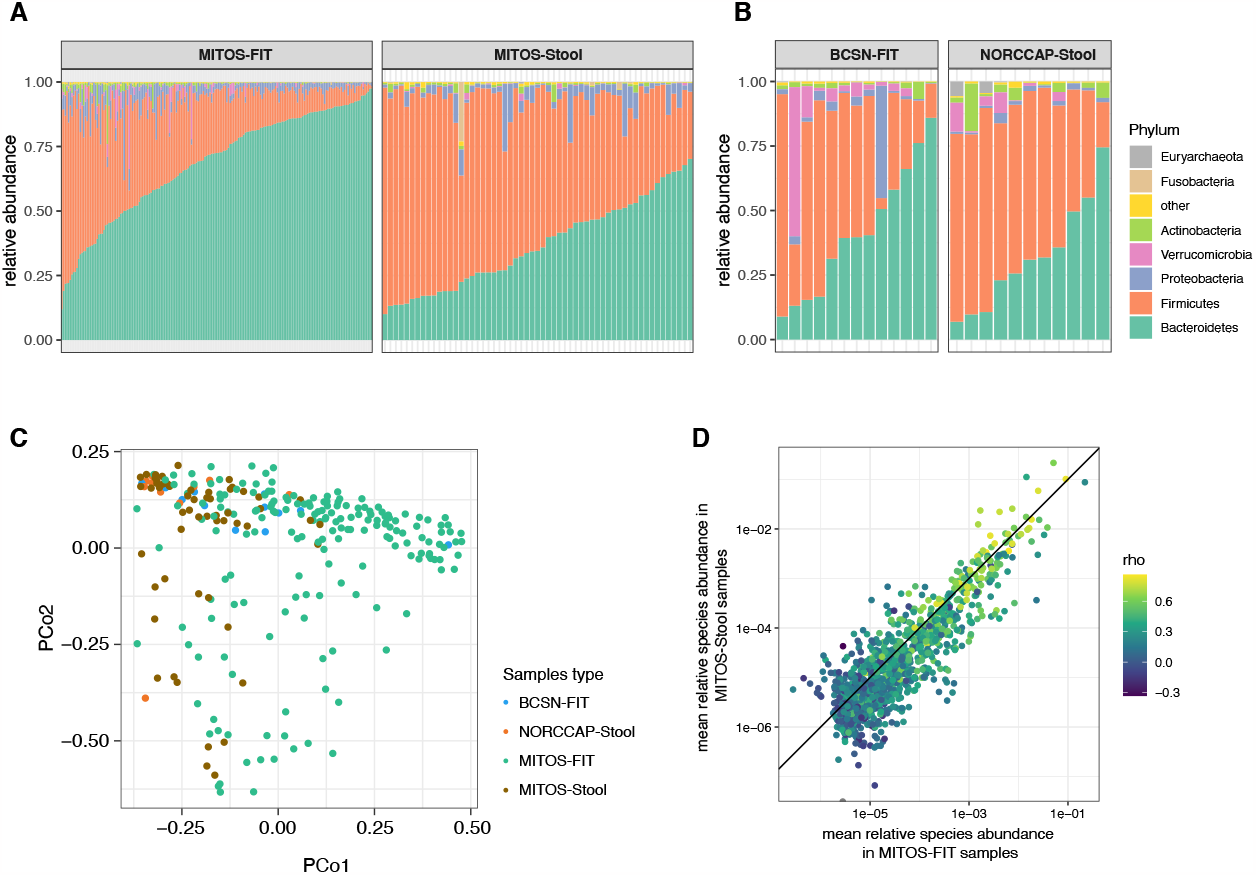
**A-B**. Classification of reads according to microbial taxa. Relative abundance of phyla in FIT-leftover and stool samples from MITOS (**A**) and NOR (**B**) cohorts. **C**. Principal coordinate (PCo) plot of FIT and stool samples (including unpaired samples). **D**. Mean relative abundance of species in stool samples (x-axis) and FIT samples (y-axis), coloured according to the Spearman’s rho of the correlation between the relative abundances in paired FIT and stool samples. The solid diagonal line represents equal abundance in either biospecimen type.

## Conclusions

Taken together, our results show that by using small RNA sequencing we can profile both stool miRNAs and microbial taxa in the left-over FIT buffer used in CRC screening. The consistent levels of miRNAs between sampling methods suggest that FIT may be used for miRNA biomarker research in large scale screening settings. This feasibility study also confirms that the alterations in gut miRNA levels in CRC patients observed in FIT samples may be used to detect miRNAs in FIT as biomarkers to improve screening performance.

## Data Availability

The datasets used and/or analysed during the current study are available from the corresponding author on reasonable request.

## Ethics approval and consent to participate

For the MITOS cohort the study design and the informed consent were approved by the local Ethics committee (AOU Città della salute e della Scienza di Torino, Italy). For the NORCCAP and BCSN cohorts anonymized samples were used. This was approved by the Norwegian Regional Ethics Committee - South-East (REC) projects, S-98052 and 2011/1272, respectively.

## Authors’ contributions

Concept and design (TBR, AN);

Collection and assembly of the data (GH, TBR, AN, CS);

Experimental tasks, data analysis and interpretation (EB, GF, BP, SUU);

Provisions of study materials or patients (GH, CS, SB);

Manuscript writing (EB, GF, BP, TBR, and AN); Manuscript editing (All);

Final approval of the version (All).

## Acknowledgements

We would like to thank Jan Inge Nordby for performing laboratory analyses for the Norwegian samples and the sequencing service was provided by the Norwegian Sequencing Centre (https://www.sequencing.uio.no), a national technology platform hosted by Oslo University Hospital and the University of Oslo supported by the Research Council of Norway and the Southeastern Regional Health Authority.

